# Metabolomics in juvenile-onset SLE: identifying new biomarkers to predict cardiovascular risk

**DOI:** 10.1101/19000356

**Authors:** George A Robinson, Kirsty E Waddington, Leda Coelewij, Ania Radziszewska, Chris Wincup, Hannah Peckham, David A Isenberg, Yiannis Ioannou, Coziana Ciurtin, Ines Pineda-Torra, Elizabeth C Jury

## Abstract

**BACKGROUND:** Juvenile-onset systemic lupus erythematosus (JSLE) is an autoimmune disorder characterised by immune dysregulation, chronic inflammation and increased cardiovascular risk (CVR). Cardiovascular disease is the leading cause of mortality in JSLE patients not attributable to disease flares. However, it is not possible to predict those patients at greatest risk using traditional CVR factors.

**METHODS:** Serum metabolomic analysis was performed using a nuclear magnetic resonance spectroscopy-platform in 31 JSLE patients. Data was analysed using cluster, linear regression and receiver operating characteristic analysis. Results were validated in a second cohort of 31 JSLE patients and using data from a cohort of adult-onset SLE patients with known pre-clinical atherosclerotic plaque.

**RESULTS:** Unbiased hierarchical clustering of metabolomic data identified three patient groups. Group-1 had decreased atheroprotective high density lipoproteins (HDL) and increased atherogenic very low and low density lipoproteins (VLDL/LDL); Group-2 had elevated HDL but reduced VLDL/LDL; and Group-3 had low HDL/VLDL/LDL levels. Notably, apolipoprotein(Apo)B1:ApoA1 ratio, a known CVR marker in adult cohorts, was elevated in Group-1 JSLE patients compared to Groups-2/3. The metabolomic signature was validated in a second JSLE cohort and compared with lipid biomarkers previously associated with pre-clinical atherosclerotic plaque in adult SLE patients. Linear regression analysis accounting for demographics, treatment, disease activity, lupus serological markers and body mass index confirmed that a unique metabolomic profile could differentiate between JSLE patients at high and low CVR.

**CONCLUSIONS:** Patient stratification using ApoB:ApoA1 ratio and lipoprotein signatures could facilitate tailored lipid modification therapies and/or diet/lifestyle interventions to combat increased CVR in JSLE.

**Key messages:**

- *What is already known about the subject?* Cardiovascular disease is the leading cause of mortality in juvenile-onset systemic lupus erythematosus (JSLE) not attributable to lupus flares; the cardiovascular risk of JSLE patients is 300 times higher than age matched healthy individuals. It is not possible to predict those patients at greatest risk using traditional risk factors.
- *What does this study add?* In depth lipoprotein-based metabolomic analysis identified Apolipoprotein(Apo)B :ApoA1 ratio as a potential biomarker for predicting increased cardiovascular risk in JSLE. This was validated in a second patient cohort and using metabolic signatures associated with pre-clinical atherosclerotic plaque development in adult SLE patients.
- *How might this impact on clinical practice or future developments?* Predicting cardiovascular risk in young JSLE patients using ApoB:ApoA1 ratio could help to stratify patients and identify those who would benefit the most from existing lipid targeting therapies. Reducing cardiovascular risk at a young age could improve patient’s life expectancy and quality of life and reduce cardiovascular comorbidity in later life.

## INTRODUCTION

Systemic lupus erythematosus (SLE) is a complex autoimmune disorder characterised by loss of immune cell regulation, chronic inflammation and multiple organ damage. Juvenile-onset disease (JSLE) (onset before 18 years) is seen in up to 20% of patients and is more severe compared to adult-onset SLE[1, 2]. Mortality from SLE has improved dramatically over the last 50 years due mainly to improved treatment[3]. However, while deaths attributable to active lupus are reduced, deaths associated with comorbidities including cardiovascular disease (CVD) are higher[4, 5]. Cardiovascular risk (CVR) and mortality are particularly increased in JSLE and it is proposed that all patients with SLE, irrespective of age at disease onset, should receive aggressive monitoring and treatment of modifiable CVR factors[6]. Currently, there are no guidelines for CVR monitoring or management in SLE/JSLE, thus there is an urgent need to find better ways to stratify patients based on their CVR and identify adequate therapeutic approaches to decrease the overall CV morbidity and mortality associated with SLE.

Atherosclerosis, a chronic inflammation of the large arteries associated with defects in lipid homeostasis, is a major cause of CVD. In patients with JSLE the atherosclerotic process begins in childhood and is accelerated in part due to prolonged exposure to inflammation[7]. Dyslipidaemia, a conventional risk factor for CVD, is a common feature of patients with both adult and JSLE, and includes elevated triglycerides (TG) and low density lipoprotein (LDL), and depressed high-density lipoprotein (HDL) and apolipoprotein-A1 (ApoA1)[8-11]. However, beyond these findings, very little information is available to assess whether defects in lipid metabolism are present in JSLE.

Interplay between traditional CVR factors (including dyslipidaemia) and risk factors associated with continuing SLE disease could contribute to development of early atherosclerosis in JSLE patients[12]. This raises the possibility that drugs targeting lipid metabolism could be beneficial in patients with JSLE. Cholesterol lowering drugs (such as statins) have been trialled in SLE patients but with mixed outcomes. Some studies show beneficial effects including improved lipid and inflammatory cytokine levels, reduced vascular inflammation, mortality and morbidity in SLE patients[13-15]. However, in two major randomised control trials, the Lupus Atherosclerosis Prevention Study (LAPS)[16] in adults and the Atherosclerosis Prevention in Paediatric Lupus Erythematosus (APPLE) trial in children[17], statins failed to meet their primary outcomes. We hypothesised that patient heterogeneity could contribute to these disappointing results and that the success of future trials could depend on the correct stratification of patients before inclusion into such studies[18, 19].

Here, in depth lipoprotein-based metabolomics was used to better characterise dyslipdaemia in patients with JSLE. Two distinct patient groups were defined and validated based on their lipoprotein profiles: Group-1 had higher levels of very low density lipoprotein (VLDL), intermediate density lipoprotein (IDL) and LDL (associated with high CVD risk), and Group-2 had higher HDL (associated with low CVD risk)[20]. Importantly, elevated apolipoprotein-(Apo)B:ApoA1 ratio was identified as a potential predictive biomarker associated with atherogenesis, this was independent of other factors including disease activity, treatment and body mass index (BMI). Collectively, these findings could help identify JSLE patients at greatest CVR and pinpoint those patients who would most benefit from targeted lipid modifying therapies.

## PATIENTS AND METHODS

### Patients

Peripheral blood was collected from JSLE patients fulfilling The American College of Rheumatology (ACR) classification criteria for lupus (1997)[21] or the Systemic Lupus International Collaborating Clinics (SLICC) criteria (2012)[22], attending a young adult or adolescent rheumatology clinic at University College London Hospital (UCLH). Two cohorts were collected: a discovery cohort (n=31) and a validation cohort (n=31). Detailed patient and disease characteristics (including demographics, disease duration, clinical manifestations and medication) were recorded from medical records and through questionnaires (Table 1). Disease activity was calculated using the SLE Disease Activity Index (SLEDAI). A score of 6 or more was used to indicate active disease[23]. Written, informed consent was acquired from all patients under the ethical approval (REC11/LO/0330). All information was stored as anonymised data. Patients treated with rituximab within the past year were excluded from the study due to previously reported substantial improvements in CVR measures following effective treatment[24].

**Table 1:** Demographic and clinical table of discovery and validation JSLE patient cohorts. For patients the SLE Disease Activity Index (SLEDAI) was calculated, a score greater than 6 represents active disease[23]. Fisher’s exact test or unpaired t-test* was used. Significant p values displayed in red. Normal ranges for lipid measures are relevant for healthy adults[54]. Abbreviations: NR: Normal ranges, BMI: Body mass index, SLEDAI: Systemic Lupus Erythematosus Disease Activity Index, ENA: Extractable nuclear antigens, Anti-CL: Anti-cardiolipin, ESR: Erythrocyte sedimentation rate, RF: Rheumatoid factor, LA: Lupus Anticoagulant, dsDNA: Anti-double-stranded-DNA antibodies, CRP: C-reactive protein, C3: Complement component 3, LC: Lymphocyte count, HDL-C: High density lipoprotein cholesterol, LDL-C: Low density lipoprotein cholesterol.

| Table 1 | JSLE (discovery) | JSLE (validation) | p-value |
| --- | --- | --- | --- |
| Total number | 31 | 31 | - |
| Female:Male | 21:10 | 30:1 | 0.0057 |
| Age, mean (range) | 19 (14-25) | 19 (13-24) | 0.8646* |
| BMI | 21.51 (20.27- 26.46) | 23.06 (20.44- 25.88) | 0.9406* |
| **Ethnicity, number (%):** |  |  |  |
| White | 11 (35%) | 8 (26%) | 0.5824 |
| Asian | 11 (36%) | 9 (29%) | 0.7863 |
| Black | 7 (23%) | 11 (35%) | 0.4018 |
| Other/unknown | 2 (6%) | 3 (10%) | >0.9999 |
| **Disease characteristics** |  |  |  |
| Age of diagnosis, mean (range) | 11.7 (0-18) | 13 (8-18) | 0.1806* |
| Disease duration, mean (range) | 7.5 (0-21) | 6.4 (0-14) | 0.3014* |
| SLEDAI, median (IQR) | 2 (0-4) | 0 (0-2) | 0.0063* |
| SLEDAI ≥6, number (%) | 7 (23%) | 1 (3%) | 0.0529 |
| SLEDAI $<$6, number (%) | 24 (77%) | 30 (97%) | 0.0529 |
| **Current organ involvement, n (%):** |  |  |  |
| Neurological | 2 (6.45%) | 9 (29%) | 0.0426 |
| Serositis | 7 (23%) | 0 (0%) | 0.0107 |
| Cutaneous | 30 (97%) | 25 (81%) | 0.1035 |
| Haematological | 13 (42%) | 12 (39%) | >0.9999 |
| Musculoskeletal | 26 (84%) | 24 (77%) | 0.7490 |
| Renal | 9 (29%) | 9 (29%) | >0.9999 |
| **Serology [median (IQR)]:** |  |  |  |
| dsDNA (IU/mL) (NR=$<$50) | 53.5 (3-332) | 21 (6-60) | 0.2694* |
| Positive ENA (number, %) | 18 (58%) | 18 (58%) | >0.9999 |
| Anti-CL IgM (MPL) (NR=0-10) | 2.7 (1.1-4.2) | 1.75 (1.1-3.93) | 0.8861* |
| Anti-CL IgG (GPL) (NR=0-20) | 1.7 (0.6-4) | 1.2 (0.35-1.9) | 0.0842* |
| CRP (mg/L) (NR$<$10) | 1 (0.6-2.58) | 0.7 (0.6-1.63) | 0.6391* |
| C3 (g/L) (NR=0.9-1.8) (mean (range)) | 0.97 (0.33-1.64) | 0.94 (0.57-1.47) | 0.6744* |
| LC (10<sup>9</sup>/L) (NR=1.3-3.5) | 1.47 (1.2-2.09) | 1.08 (0.91-1.25) | 0.0020* |
| **Clinical lipids [median (IQR)]:** |  |  |  |
| Cholesterol (NR$<$5mmol/L) | 3.95 (3.48-4.2) | 4 (3.4-4.9) | 0.9883* |
| Triglycerides (NR$<$3mmol/L) | 0.8 (0.58-1.23) | 0.8 (0.5-1) | 0.8038* |
| HDL-C (NR$>$1mmol/L) | 1.45 (1.25-1.7) | 1.4 (1.1-1.7) | 0.8764* |
| LDL-C (NR$<$3mmol/L) | 2 (1.6-2.23) | 2.2 (1.5-2.6) | 0.9351* |
| Cholesterol:HDL (NR$<$4) | 2.55 (2.2-3.35) | 2.9 (2.5-3.4) | 0.7889* |
| **Current treatment [n (%]):** |  |  |  |
| Hydroxychloroquine | 27 (87%) | 29 (94%) | 0.6713 |
| Mycophenolate mofetil | 15 (48%) | 9 (29%) | 0.1919 |
| Prednisolone | 16 (52%) | 11 (35%) | 0.3056 |
| Vitamin D | 7 (23%) | 6 (19%) | >0.9999 |
| Methotrexate | 3 (10%) | 4 (13%) | >0.9999 |
| Azathioprine | 7 (23%) | 8 (26%) | >0.9999 |
| **Past rituximab treatment (%)** |  |  |  |
| Rituximab in the last year | 0 (0%) | 0 (0%) | >0.9999 |
| Rituximab ever | 3 (8.6%) | 1 (1%) | 0.2906 |

### Patient and public involvement

Patients were not involved with the design of the study. However, using online surveys[25] and face-to-face events[26] patients have been consulted about their experience of CVR counselling and management, the acceptability of potential therapeutic interventions and the design of future clinical trials.

### Metabolomics

Serum metabolomics analysis was performed using nuclear magnetic resonance (NMR) spectroscopy by Nightingale Health (https://nightingalehealth.com/)[27] (Supplementary Table 1 for list of metabolites).

### Statistical analysis

Statistical analysis was performed using GraphPad Prism-7. Data was tested for normal distribution using Kolmogorov-Smirnov test and parametric/non-parametric tests were used accordingly. Unpaired and paired t-tests and One-way ANOVA were used as appropriate. Linear regression was performed using a 95% confidence interval to calculate significance (Pearson correlation). Multiple testing was accounted for using the Holm-Sidak correction of p-values. Receiver operating characteristic (ROC) curve analysis was performed using GraphPad Prism-7.

MultiExperiment Viewer (MeV) was used to produce heat maps and hierarchical clustering (Pearson correlation) using z-score transformed raw data values calculated in Microsoft Excel 2010 using the equation: *((Individual Value-Population Mean)/(Population Standard Deviation–Square Root of the Total Sample Number))*.

ClustVis web-tool[28] was used to perform principle component analysis.

Logistic regression was performed in RStudio[29] on each metabolomic biomarker and adjusted for demographic, clinical and treatment parameters. Results were visualised using the R package foresplotNMR[30].

## RESULTS

### Stratification of JSLE patients using detailed lipoprotein profiles identified three unique patient groups

Dyslipidaemia was investigated in patients with JSLE using an in-depth metabolomics platform (Discovery cohort, Table-1, and Supplementary Table 1 for metabolite list). Lipoprotein measures were converted to z-scores and unbiased hierarchical clustering was performed (Figure 1A). Despite routine clinical serum lipid measures being largely within normal ranges (Table-1), three distinct clusters based on lipoprotein particle concentrations and diameters were identified: Group-1 characterised by reduced serum HDL and elevated VLDL/IDL/LDL particles; Group-2 with elevated HDL and reduced VLDL/IDL/LDL particles and; Group-3 had relatively low serum levels of HDL and VLDL/LDL/IDL particles (Figure 1A and B) Beyond lipoproteins, other lipid metabolites showed a distinct pattern of distribution within the three clusters (Figure 1C). Patients in Group-1 (high VLDL/IDL/LDL) were characterised by increased levels of total, free and esterified VLDL/IDL/LDL-associated cholesterol, free and lipoprotein-associated TG levels, choline related lipids (phosphoglycerides (PG), phosphatidylcholine (PC) and sphingomyelins (SM)) and polyunsaturated, omega and saturated fatty acids (FA) whereas HDL cholesterol and unsaturated FAs were reduced compared to the overall cohort average. The opposite was found in Groups-2 and 3 where VLDL/IDL/LDL-associated cholesterol and TG levels were lower in all cases. Groups-2 and 3 had a similar metabolomic profile except that JSLE patients in Group-3 had additionally lower HDL levels (Figure 1B). Thus, three distinct JSLE patient groups could be stratified based on their metabolomic lipoprotein profile alone.

**Figure 1:**
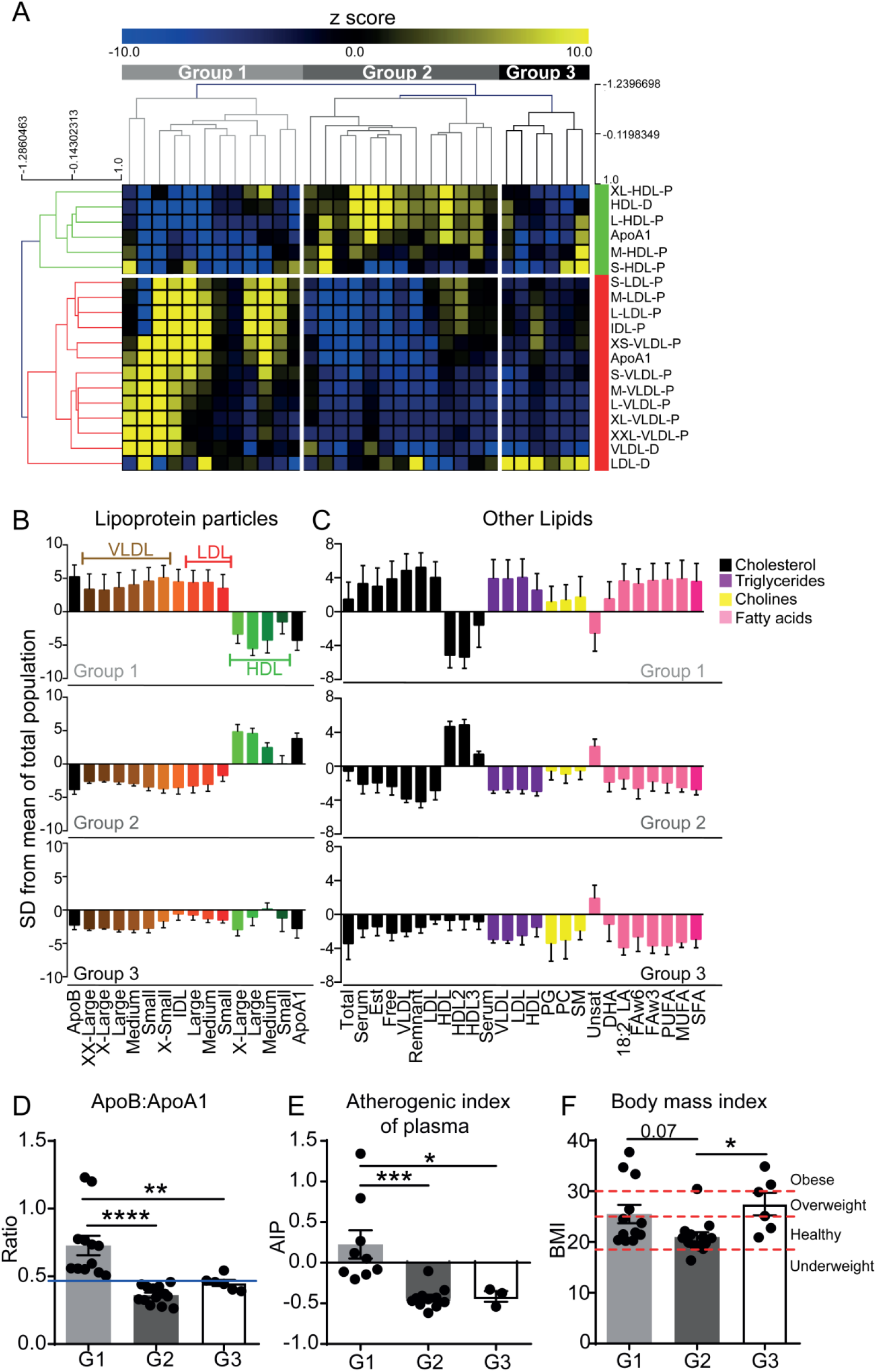
JSLE patient stratification by lipoprotein profile. Metabolomics analysis was performed on serum from 31 JSLE patients (Discovery cohort). **(A)** Heat map displaying the z-score converted measurements of lipoprotein particle (P) concentrations and diameters (D) and apolipoproteins (Apo). Unbiased hierarchical clustering was performed using MeV software and 3 groups are labelled (Group-1=light grey, Group-2=dark grey, Group-3=black). Anti- and pro-atherogenic lipoproteins clustered (labelled in green and red respectively). **(B)** Measurements from each stratified group showing lipoprotein subclasses (as shown in (A)) and **(C)** other lipid phenotypes. Data in units of standard deviation (SD) are shown as deviation from the mean value of the whole study population. Error bars indicate standard error. Abbreviations: Apo, apolipoprotein, VLDL, IDL, LDL, HDL, very low, intermediate, low and high density lipoproteins, XX-large, chylomicrons and extremely large; X-Large, very large; X-small, very small; Est (esterified), PG (Phosphoglyceride), PC, Phosphatidylcholine; SM, Sphingomyelins; Unsat, Unsaturated; DHA, Docosahexaenoic acid; LA, Linoleic acid; FAw3, Omega-3 fatty acids; FAw6, Omega-6 fatty acids; PUFA, Polyunsaturated fatty acids; MUFA, Monounsaturated fatty acids; SFA, Saturated fatty acids. Assessment of cardiovascular risk measures across three patient groups: Group-1 (n=12), Group-2 (n=13) and Group-3 (n=6). **(D)** ApoB:ApoA1 ratio. Mean, One-way ANOVA (**=P<0.01, ****=P<0.0001). Red line represents the mean ApoB:ApoA1 ratio of age and sex matched healthy controls (n=32; 15 males, 17 females). **(E)** Atherogenic index of plasma [Log(Triglycerides/HDL-Cholesterol)]. Mean, One-way ANOVA, (*=P<0.05, ***=P<0.001). **(F)** Body Mass Index (BMI) of patients. Dashed red lines indicate the cut off BMI values for underweight, healthy, overweight and obese. Mean, One-way ANOVA. (*=P<0.05).

### JSLE patients in the three metabolic groups had a similar clinical profile but were associated with an altered CVR

Strikingly, the clinical and disease features of JSLE patients in each of the three metabolic groups were similar (Supplementary Table 2). There were no significant differences in disease activity score (SLEDAI), age of onset, disease duration or treatment. Patients in Group-1 had significantly reduced complement C3 compared to patients in Group-3. Notably, although almost all patients had total cholesterol and TG levels within normal ranges, patients in Group-1 had reduced HDL cholesterol and elevated LDL cholesterol compared to Groups-2 and −3 (Supplementary Table 2).

Recent studies show that the ratio between serum ApoB:ApoA1 is a more effective CVR predictor than routine cholesterol measurements; a higher ratio is associated with increased risk[31, 32]. JSLE patients in Group-1 had a significantly elevated ApoB:ApoA1 ratio compared to Groups-2 and 3 (Figure 1D); this was higher than the mean value from age matched adolescent healthy donors. In support of this finding, other measures associated with increased CVR were also altered between the three groups; patients in Group-1 had a significant increase in atherogenic index of plasma (Log(Triglycerides/HDL-Cholesterol)[33]) compared to Groups-2 and 3 (Figure 1E) and BMI was significantly higher in patients in Group-3 compared to Group-2 (Figure 1F).

### Metabolomic stratification was validated and associated with markers of pre-clinical atherosclerotic plaque

The robustness of this metabolomic stratification was tested in a second cohort of 31 JSLE patients, recruited with no selection bias other than no rituximab treatment in the previous year (Validation cohort, Table 1). Using the same lipoprotein markers and unbiased hierarchical clustering method, patients clustered into 2 groups (Figure 2A). These 2 groups (labelled Group-1A and 2A) reflected the lipoprotein phenotype observed in Groups-1 and 2 in the discovery cohort (Figure 1A). Group-3 seen in the discovery cohort was not validated. Importantly, measures associated with CVR in the general population were significantly increased in JSLE Group-1A including the ApoB:ApoA1 ratio, atherogenic index of plasma and BMI compared to Group-2A (Figure 2B-D). As in the discovery cohort, no major clinical parameters discriminated between Groups-1A and 2A (Supplemental Table 3).

**Figure 2:**
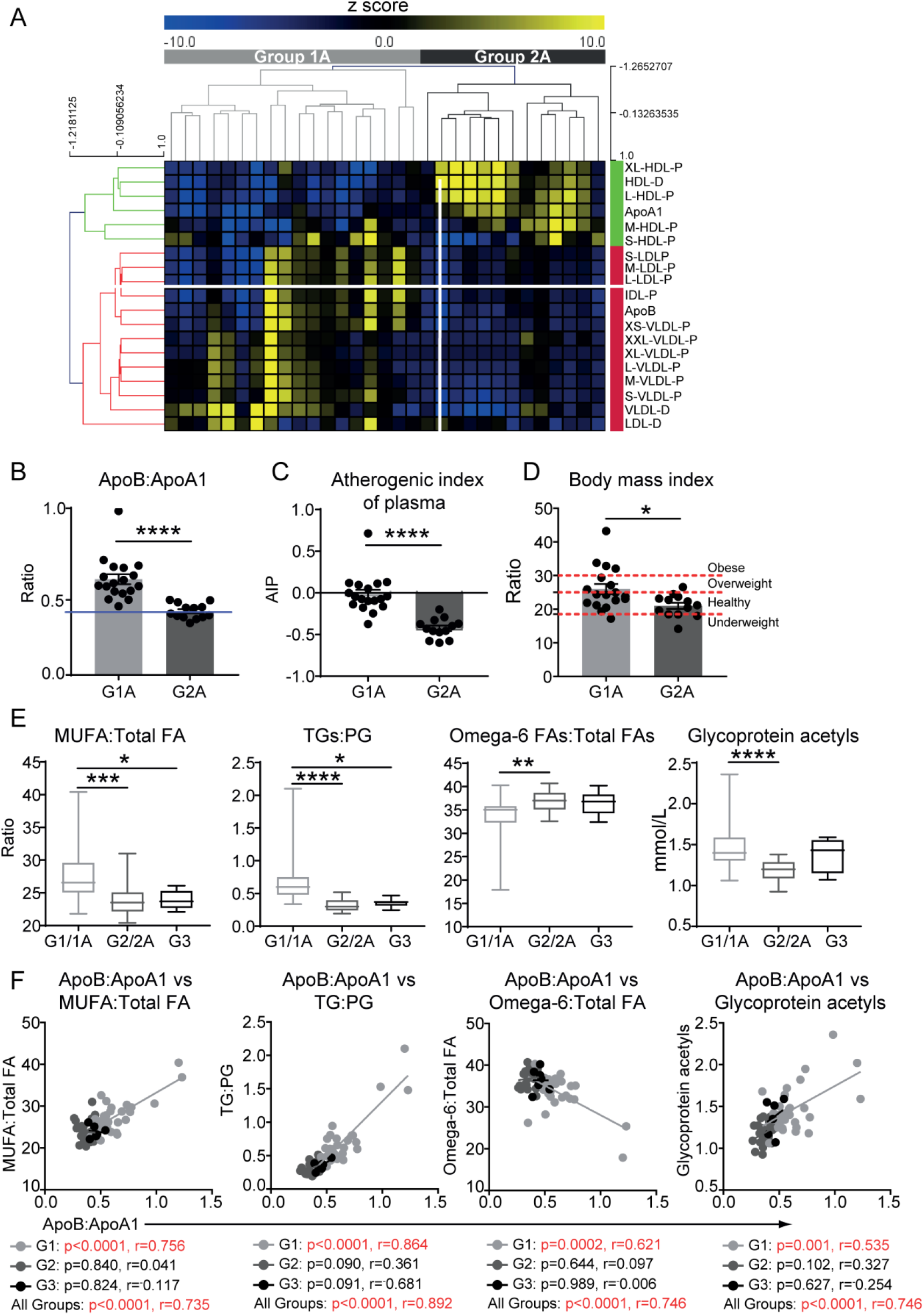
Validation of cardiovascular risk groups in JSLE. Metabolomics analysis was performed on serum from an additional 31 JSLE patients (Validation cohort). **(A)** Heat map displaying the z-score converted measurements of lipoproteins and apolipoproteins. Unbiased hierarchical clustering was performed; 2 groups are labelled (Group-1A=light grey and Group-2A=dark grey. Anti- and pro-atherogenic lipoproteins clustered (labelled in green and red respectively). Assessment of cardiovascular risk measures across validated patient groups: Group-1A (n=18) and Group-2A (n=13). **(B)** ApoB:ApoA1 ratio. Red line represents the mean ApoB:ApoA1 ratio of age and sex matched healthy controls (n=17; 17 females). Mean, t-test, ****=P<0.0001. **(C)** Atherogenic index of plasma [Log(Triglycerides/HDL-Cholesterol)]. Mean, t-test, ****=P<0.0001 **(D)** Body Mass Index (BMI) of patients. Dashed red lines indicate the cut off BMI values for underweight, healthy, overweight and obese. Mean, t-test, *=P<0.05. **(E)** Levels of four metabolites we identified previously associated with pre-clinical plaque in adult SLE patients[34]. Discovery and validation JSLE cohort data were combined (Group1+1A (n=30), Group 2+2A (n=26)) and discovery cohort Group 3 (n=6). One-way ANOVA, *=P<0.05, **=P<0.01, ***=P<0.001, ****=P<0.0001. **(F)** Correlations between serum ApoB:ApoA1 levels and biomarkers associated with pre-clinical atherosclerotic plaque in adult SLE patients[34] using combined data from the discovery and validated JSLE groups (Group1+1A (n=30, light grey), Group 2+2A (n=26, dark grey) and discovery cohort Group 3 (n=6, black). Pearson’s correlation coefficient (r) and significance were determined using a 95% confidence interval. Statistically significant p values are displayed in red. FA: Fatty acids, MUFA: Monounsaturated fatty acids, TG: Triglycerides, PG: Phosphoglycerides.

In order to confirm that JSLE patients in Groups-1/1A had an atherogenic lipid profile, a comparison was made with metabolites previously identified to be associated with pre-clinical carotid and femoral artery atherosclerotic plaque in adult-onset SLE patients[34]. Lipid biomarkers identified in adult SLE patients with pre-clinical plaque - elevated ratios of triglycerides:phosphoglycerides and monounsaturated:total fatty acids, elevated glycoprotein acetyl levels and reduced ratios of omega-6:total fatty acids - were also significantly altered in Group-1/1A JSLE patients compared to Groups-2/2A and 3 (Figure 2E). Furthermore, these plaque-associated metabolites correlated significantly with the ApoB:ApoA1 ratio in patients from Group1+1A only (Figure 2F).

Together, these results suggested that metabolomic analysis could stratify JSLE patients according to their CVR.

### ApoB:ApoA1 ratio could be an independent biomarker associated with increased CVR in patients with JSLE

In order to further assess the utility of specific biomarkers in predicting CVR in JSLE patients, logistic regression analysis was performed on the data comparing Groups-1+1A with Groups-2+2A; data was normalised for age, sex, ethnicity, BMI, disease duration, disease activity, double-stranded DNA autoantibodies, C-reactive protein, complement-C3, lymphocyte count and treatment (Figure 3, Supplementary Figure 1, Supplementary Data Table 1). ApoB:ApoA1 ratio was confirmed as a biomarker of CVR independent from disease and demographic features. In addition, a number of other lipid metabolites were identified including VLDL, IDL and LDL lipid content and monounsaturated fatty acids associated with the high CVR group whereas HDL lipid content and unsaturated fatty acids were associated with low CVR group (Figure 3). Interestingly Glycoprotein Acetyl, related with increased cardiovascular mortality risk [35, 36], was the only non-lipid metabolite differentially expressed between the groups (Supplementary Figure 1). In order to further assess the power of the identified biomarkers, stringent p value correction for multiple testing was applied to the logistic regression data; 11 metabolites were differentially associated with potential high and low CVR in JSLE (Figure 4A, Supplementary Table 4). This included increased ApoB:ApoA1 ratio and concentration of small VLDL cholesterol esters (VLDL-CE) associated with high CVR and reduced large HDL particle concentration, lipid content and diameter associated with low CVR. Notably, the majority of these metabolites (Figure 4A) can be significantly affected by statins[37, 38], pro-protein convertase subtilisin/kexin type 9 (PCSK9) inhibitors[39], dietary fish consumption and/or BMI[40]. Principle component analysis using the top 11 differentially expressed metabolites between Group1+1A and Group2+2A confirmed independent clustering of the 2 groups (Figure 4B).

**Figure 3:**
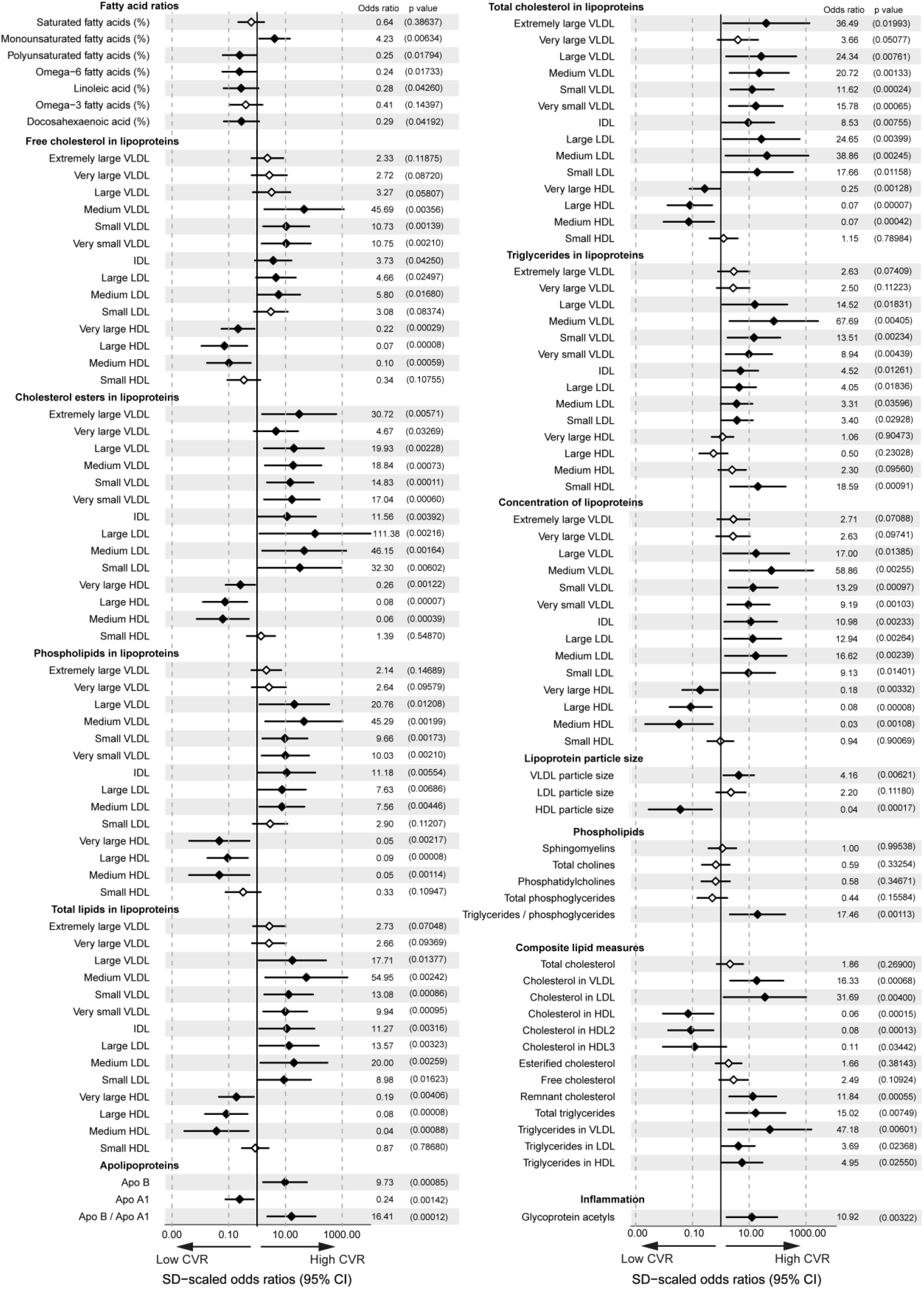
Odds ratios of lipid and inflammatory metabolites between high and low CVR groups adjusting for clinical parameters. Forest plot displaying the odds ratios of blood lipid and inflammatory metabolites between Group1+1A (high CVR risk, n=30) and Group2+2A (low CVR risk, n=26) adjusted for age, sex, ethnicity, treatments, BMI and disease parameters. Significances between groups are displayed as a filled in point with p values displayed in brackets to the right. Metabolites significantly altered by clinical and demographic parameters are shown in Supplementary Data Table 1.

**Figure 4:**
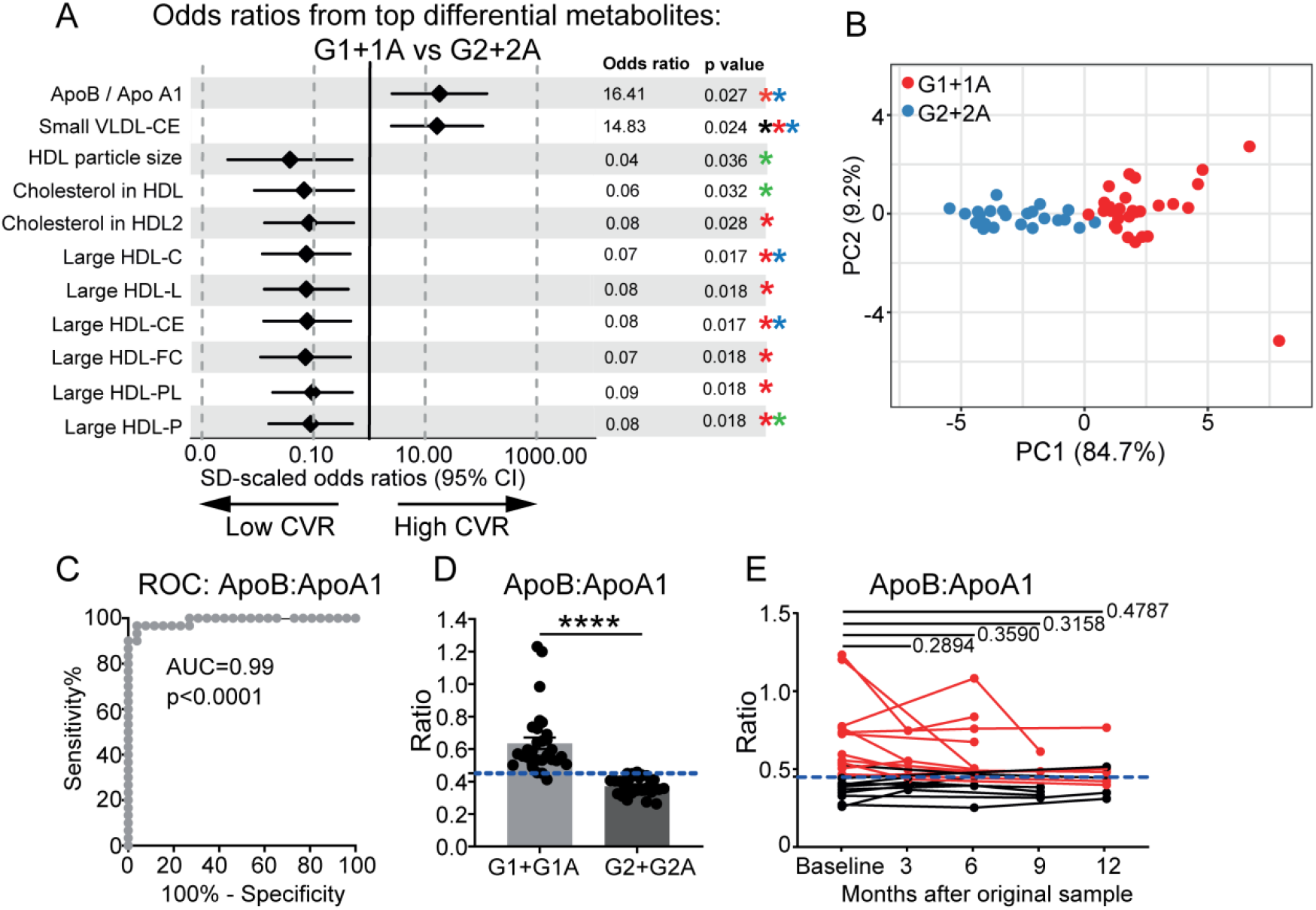
ApoB:ApoA1 as a clinical biomarker of CVR in JSLE. (**A**) Forest plot from linear regression analysis displaying the odds ratio and 95% CI of 11 metabolites that passed p value correction for multiple testing between Group1+1A (n=30) and Group2+2A (n=26). Data was normalised for clinical parameters, demographic information, treatment and BMI. Corrected p-values (Holm-Sidak) are displayed for each metabolite. Stars represent metabolites that are significantly affected by BMI (Black), statins (Red)[37-39], PCSK9 inhibitors (Blue)[39] and habitual dietary fish consumption (Green)[40]. (**B**) Principle component analysis using the 11 significant metabolites between Group1+1A (n=30, red) and Group2+2A (n=26, blue) identified in (A). **C)** ROC curve analysis of the ApoB:ApoA1 ratio of patients in Group-1+1A (n=30) compared to patients in Group-2+2A (n=26). Area under the curve (AUC) is displayed. **D)** ApoB:ApoA1 ratio (higher ratios=higher risk) measured in Group-1+1A (n=30) and Group-2+2A (n=26). Cut-off identified from the ROC analysis is displayed as the blue dashed line. Mean, unpaired two-tailed t-tests, ****=P<0.0001. **E)** Longitudinal analysis of ApoB:ApoA1 ratio at 3 month intervals over 12 months (baseline (n=22), 3 months (n=9), 6 months (n=12), 9 months (n=6) and 12 months (n=9)). The cut off identified from the ROC curve analysis is displayed as the blue dashed line Paired t-tests.

Finally, a ROC curve analysis of the 11 differentially expressed metabolites confirmed that ApoB:ApoA1 ratio had a highly significant predictive capacity to distinguish patients in the high CVR Groups-1+1A from patients in low CVR Groups-2+2A (area under the curve=0.99, p<0.0001) (Figure 4C and Supplementary Figure 2A). From this analysis a highly sensitive and specific cut-off ApoB:ApoA1 ratio of 0.45 was established to enable these groups to be distinguished from one another (Figure 4D). Importantly, routine clinical measures of lipids were less effective at distinguishing between the groups (Supplementary Figure 2B). Longitudinal analysis of ApoB:ApoA1 ratios from repeat patient visits (3 month intervals) over a 12 month period showed that the ApoB:ApoA1 ratio biomarker remained stable over time (Figure 4E) despite individual patient fluctuations in disease activity scores (Supplementary Figure 3).

Therefore, the results suggest that ApoB:ApoA1 ratio is a robust biomarker that could be used to predict increased CVR in patients with JSLE. These high risk patients could not be confidently distinguished using routine clinical assessment or standard lipid measures. Lipid modification therapies and/or diet could be beneficial to modify CVR in patients with a high ApoB:ApoA1 ratio (Figure 5).

**Figure 5:**
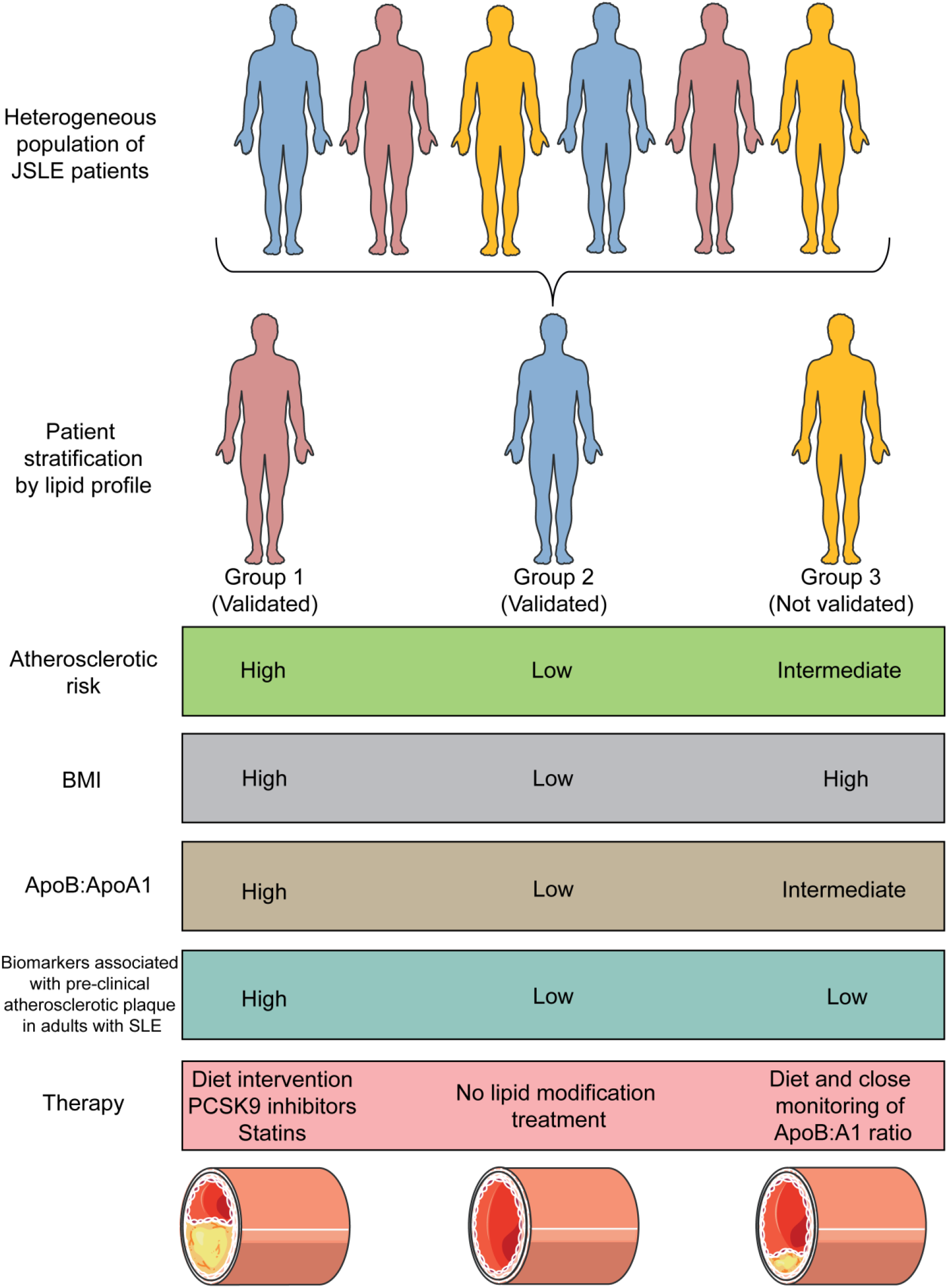
Phenotypic summary of the stratified JSLE patient groups. Summary of atherosclerotic risk and proposed therapeutic options for lipid stratified JSLE patient groups. Therapeutic options include diet and/or therapies shown to be effective in large randomised trials in lowering cardiovascular disease risk. Abbreviations: Apo (Apolipoprotein), BMI (Body Mass Index), PCSK9 (Pro-protein convertase subtilisin/kexin type 9). Images were provided from Servier Medical Art, licensed under a Creative Common Attribution 3.0 Generic License http://smart.servier.com.

## Discussion

JSLE patients have an increased risk of developing CVD. Despite this knowledge, CVD is still the leading comorbidity for patients[2, 5]. This study used detailed serum metabolomics to stratify patients with JSLE into two validated groups, those with high and low CVR, based on lipoprotein particle concentrations and diameters and apolipoprotein concentrations. CVR was validated through the measurement of metabolomic markers associated with pre-clinical atherosclerotic plaque in adult SLE patients[34]. ApoB:ApoA1 was found to be a reliably predictive biomarker of CVR in JSLE irrespective of demographic features (including BMI) and disease/treatment measures. This information strongly suggests that a group of JSLE patients with a potential increased CVR could benefit from tailored lipid modifying therapy and/or diet intervention and that stratification based on more detailed lipoprotein taxonomy should be included in future clinical trials.

Prediction of cardiovascular disease in the general population predominantly relies on relatively simple measurements such as serum total cholesterol, HDL, LDL and triglycerides[20]. However over recent years is has become apparent that more in-depth lipoprotein assessment including lipoprotein subclasses and their lipid concentrations and composition can provide new insight into CVR prediction and understanding of disease mechanisms[36, 41]. Using this approach could be particularly relevant for assessing CVR in patients with chronic inflammatory conditions such as JSLE, which is difficult to assess in the absence of traditional risk factors[3]. Dyslipidaemia (assessed using routine clinical measures) is reported in JSLE, however it is difficult to differentiate the effect of disease activity and treatment on lipid profiles from conventional CVR lipid profiles[9-11].

To our knowledge, no studies have used detailed lipoprotein profiles to stratify JSLE patients to assess their CVR. Using this method we characterised ApoB:ApoA1 ratio as a potential biomarker for increased CVR in JSLE patients. This finding supports findings in the Cardiovascular Risk in the Young Finns Study showing that elevated ApoB and ApoB:ApoA1 ratio and reduced ApoA1 in children and adolescents reflected a predisposition to subclinical atherosclerosis in adulthood in healthy individuals[42] and previous reports showing that ApoB:A1 ratio has improved CVR predictive value compared to conventional blood cholesterol measures[31, 32]. Reduced ApoA1 and reduced HDL concentration have been reported previously in JSLE patients compared with healthy donors[43] and increased Apo-B was associated with arterial stiffness in adult SLE patients[44].

A cut-off value for ApoB:ApoA1 ratio of 0.45 was identified that could have clinical utility. Comparison to other published work showed that ApoB:ApoA1 ratio is age dependant so further validation of age specific cut points is needed. For example, a study of healthy and overweight adults (mean age 56 years) reported an ApoB:ApoA1 ratio of 0.80 as predictive of coronary heart disease in overweight/obese people, and that this performed better than other traditional clinical measures[36]. Alternatively, a Swedish study defined a cut-off of 0.63 for femoral artery atherosclerosis prediction in women (mean age 64-years)[45]. Another study, including young healthy females identified ApoB:A1 ratios from 0.55 at age 10-years to 0.58 at age 19-years [46]. Differences in published ratios could also be accounted for by different technologies used to assess ApoB and ApoA1 levels (mainly colorimetric methodology compared to the NMR technology used here) and the fasting status of the study participants (fasting versus non-fasting); our study was performed on non-fasting patients.

Not all patients with JSLE have the same CVR; therefore identifying low and high CVR groups in clinic could be pivotal for the success of interventions to reduce this risk. A 3-year clinical trial of atorvastatin in patients with JSLE aged 10-21 (the APPLE trial) did not meet its primary outcome of reduced progression of subclinical atherosclerosis, despite atorvastatin-treated participants showing a trend towards slower carotid intima-media thickness (CIMT) progression[17]. However, the authors suggested that certain subgroups of JSLE patients would benefit from targeted statin therapy, although the routine serum lipid level measurements did not facilitate patient stratification. This study showed that subclinical atherosclerosis in SLE begins in the paediatric age group and highlighted the need for paediatric rheumatologists to address modifiable CVR factors in daily practice. Other patient groups responding well to treatment with statins, as assessed by HDL and LDL-C levels, have been shown to have high ApoB[47, 48]; thus apolipoprotein measures could help optimise the efficacy of lipid modifying therapy by targeting the correct patients. In addition to statins other interventions could also be relevant including PCSK9 inhibitor[49] and nutritional intervention through increased dietary intake of omega-3 FAs[50, 51]. The efficacy of personalised nutritional therapy has been validated recently in patients with diabetes; high variability between post-meal glucose levels in patients was used to stratify patients for specific short-term personalised dietary interventions to manage glucose levels[52]. Dietary intervention has been shown to reduce disease activity scores in SLE patients[50, 53] and patients are very engaged with such strategies[25].

It will be important to validate this work in a larger multi-centre study to account for genetic, environmental and dietary differences, although this cohort was ethnically diverse. Future research will address both clinical validation (including endothelial dysfunction studies and measurement of plaque and intimal-media thickness) as well as potential therapeutic interventions.

In conclusion, there are no guidelines for CVR monitoring or management in patients with SLE/JSLE, thus there is an urgent need to find better ways to stratify these patients based on their CVR and identify adequate therapeutic approaches to decrease the overall CV morbidity and mortality associated with SLE. This study proposes high ApoB:ApoA1 ratio as a potential biomarker to stratify JSLE patients for a more personalised approach to lipid modification therapy.

## Data Availability

Metabolomic and logistic regression data is listed in the supplemental data.

## Contributors

Design of research study; ECJ, ITP, CC, YI. Acquiring data; GAR, KW; AR, HP, CW. Recruiting patients; HP, YI, CC, CW, DAI. Analyzing data; GAR, LC: Writing the manuscript; GAR, ECJ: Review of the manuscript; DAI, CC, ITP. All authors approved the final version.

## Ethics approval

This study was approved by the London-Harrow research Ethics Committee, 11/LO/0330

